# Long-term outcomes of hospitalized SARS-CoV-2/COVID-19 patients with and without neurological involvement: 3-year follow-up assessment

**DOI:** 10.1101/2023.06.26.23291883

**Authors:** Anna Eligulashvili, Moshe Gordon, Jimmy S Lee, Jeylin Lee, Shiv Mehrotra-Varma, Jai Mehrotra-Varma, Kevin Hsu, Imanyah Hilliard, Kristen Lee, Arleen Li, Muhammed Amir Essibayi, Judy Yee, David J Altschul, Emad Eskandar, Mark F. Mehler, Tim Q. Duong

## Abstract

**Background and Objectives:** Acute neurological manifestations are a common complication of acute COVID-19 disease. This study investigated the 3-year outcomes of patients with and without significant neurological manifestations during initial COVID-19 hospitalization.

**Methods:** Patients infected by SARS-CoV-2 between March 1 and April 16, 2020 and hospitalized in the Montefiore Health System in the Bronx, an epicenter of the early pandemic, were included. Follow-up data was captured up to January 23, 2023 (3 years post COVID-19). This cohort consisted of 414 COVID-19 patients with significant neurological manifestations and 1199 propensity-matched COVID- 19 patients without neurological manifestations. Primary outcomes were mortality, stroke, heart attack, major adverse cardiovascular events (MACE), reinfection, and hospital readmission post-discharge. Secondary outcomes were clinical neuroimaging findings (hemorrhage, active stroke, prior stroke, mass effect, and microhemorrhage, white-matter changes, microvascular disease, and volume loss). Predictive models were used to identify risk factors of mortality post-discharge.

**Results:** More patients in the neurological cohort were discharged to acute rehabilitation (10.54% vs 3.68%, p<0.0001), skilled nursing facilities (30.67% vs 20.78%, p=0.0002) and fewer to home (55.27% vs 70.21%, p<0.0001) compared to the matched controls. Incidence of readmission for any medical reason (65.70% vs 60.72%, p=0.036), stroke (6.28% vs 2.34%, p<0.0001), and MACE (20.53% vs 16.51%, p=0.032) was higher in the neurological cohort post-discharge. Neurological patients were more likely to die post-discharge (58 (14.01%) vs 94 (7.84%), p=0.0001) compared to controls (HR=2.346, 95% CI=(1.586, 3.470), p<0.0001). The major causes of death post-discharge were heart disease (14.47%), sepsis (13.82%), influenza and pneumonia (11.18%), COVID-19 (8.55%) and acute respiratory distress syndrome (7.89%). Factors associated with mortality after leaving the hospital were belonging to the neurological cohort (OR=1.802 (1.237, 2.608), p=0.002), discharge disposition (OR=1.508, 95% CI=(1.276, 1.775), p<0.0001), congestive heart failure (OR=2.281 (1.429, 3.593), p=0.0004), higher COVID-19 severity score (OR=1.177 (1.062, 1.304), p=0.002), and older age (OR=1.027 (1.010, 1.044), p=0.002). There were no group differences in gross radiological findings, except the neurological cohort showed significantly more age-adjusted brain volume loss (p<0.05) compared to controls.

**Discussion:** COVID-19 patients with neurological manifestations have worse long-term outcomes compared to matched controls. These findings raise awareness and the need for closer monitoring and timely interventions for COVID-19 patients with neurological manifestations.

## INTRODUCTION

Severe acute neurological events – such as ischemic stroke, seizure, intracranial hemorrhage and thrombosis, and encephalopathy – have been reported in COVID-19 patients(1–8). The causes of these central nervous system (CNS) manifestations are multifactorial. There is conflicting evidence whether SARS-CoV-2 infects neuronal cells, with some studies reporting neuronal invasion(9), while others report no evidence of direct infection(10, 11). Additional studies suggest that a diffuse microvasculopathy may ensue with endothelial compromise, micro-infarctions, subsequent micro-hemorrhages, and microglial conglomerates with innate immune activation. Nonetheless, CNS manifestations could also arise from secondary effects, such as from respiratory distress, cardiovascular stress, sepsis, hypercoagulation, and host-mediated immune responses triggered by SARS-CoV-2 infection. Patients with neurological complications have been shown to have worse *acute* COVID-19 outcomes compared to propensity-matched controls(12). However, the long-term outcomes of COVID-19 survivors with CNS manifestations are unknown.

The goal of this study was to evaluate the 3-year outcomes of COVID-19 patients with significant neurological complaints that warranted neuroimaging during COVID-19 when compared with propensity-matched controls without significant neurological complaints. Improved understanding of the long-term outcomes of COVID-19 patients with CNS manifestations could help to identify at-risk patients and enable timely interventions to address the potentially high burden of care among these COVID-19 survivors.

## METHODS

### Data sources

This observational study followed the STROBE guideline. This is a follow-up retrospective study of a previously reported retrospective cohort study of adult patients(12) admitted to the Montefiore Health System due to COVID-19 between March 01 and April 16, 2020 with confirmed SARS-CoV-2 infection by real-time reverse transcriptase PCR-positive assay testing. Follow-up data was captured up to January 23, 2023 (3 years follow-up).

The original neurological cohort consisted of 636 hospitalized COVID-19 patients who experienced various neurological signs and symptoms that warranted neuroimaging during COVID-19 hospitalization. Neurological involvement included acute stroke (confirmed by imaging), new or recrudescent seizures, anatomic brain lesions (subdural hematoma, brain tumor, chronic infarction, or nonspecific lesions), presence of altered mentation with evidence for impaired cognition or arousal, and neuro-COVID-19 complex (headache, anosmia, ageusia, chemesthesis, vertigo, presyncope, paresthesias, cranial nerve abnormalities, ataxia, dysautonomia, and skeletal muscle injury with normal orientation and arousal signs). The original control group consisted of 1743 hospitalized COVID-19 patients by 3:1 propensity-matching for age and COVID-19 severity score who did not have significant neurological issues during hospitalization. After excluding patients who died during hospitalization or were missing from our database, the neurological cohort and control cohort had sample sizes of 414 and 1199 patients, respectively. Note that the samples differed slightly from the original paper because a few additional patients were found to meet the inclusion/exclusion criteria.

### Data abstraction

Health data were extracted automatically from the electronic medical records as described previously(13–18). De-identified health data were obtained for research after standardization to the Observational Medical Outcomes Partnership (OMOP) Common Data Model (CDM) version 6. OMOP CDM represents healthcare data from diverse sources, which are stored in standard vocabulary concepts(19), allowing for the systematic analysis of disparate observational databases, including data from the electronic medical record (EMR), administrative claims, and disease classifications systems (e.g., ICD-10, SNOWMED, LOINC, etc.). ATLAS, a web-based tool developed by the Observational Health Data Sciences and Informatics (OHDSI) community that enables navigation of patient-level, observational data in the CDM format, was used to search vocabulary concepts and facilitate cohort building. Data were subsequently exported and queried as SQLite database files using the DB Browser for SQLite (version 3.12.0). To ensure data accuracy, our team performed extensive cross validation of all major variables extracted by manual chart reviews on subsets of patients(13–18).

### Discharge dispositions

Discharge disposition of survivors from COVID-19 hospitalization were categorized as home (with or without home care), hospice, acute rehabilitation, skilled nursing facility (SNF), and others (i.e., custodial care, supportive care, and psychiatric care).

### Data abstraction

Age, gender, race, ethnicity, comorbidities, and laboratory test data were extracted from electronic medical records at follow-up extending to January 23, 2023. Incidence of stroke, heart attack, major adverse cardiac events (MACE, defined as the composite of cardiovascular death, nonfatal stroke, nonfatal myocardial infarction, new-onset nonfatal heart failure, thromboembolism and nonfatal cardiogenic shock(20, 21)), reinfection, and readmission after COVID-19 discharge for any reason were tabulated at 3-year follow-up. For non-survivors, the cause of death was ascertained via chart review and categorized using the primary reason for death as reported on the death certificate.

Preexisting comorbidities included chronic obstructive pulmonary disease (COPD) and asthma, diabetes, congestive heart failure (CHF), and chronic kidney disease (CKD). Vital signs and laboratory data included temperature, systolic blood pressure (SBP), arterial pressure, D-dimer (DDIM), international normalized ratio (INR), blood urea nitrogen (BUN), creatinine (Cr), sodium, glucose, aspartate transaminase (AST), alanine aminotransferase (ALT), white blood cell count (WBC), lymphocyte count (Lymph) ferritin (FERR), C-reactive protein (CRP), procalcitonin, lactate dehydrogenase (LDH), brain natriuretic peptide (BNP), troponin-T (TNT), and arterial oxygen saturation. Laboratory data for COVID-19 admission and at follow-up (most recent) were obtained.

### COVID-19 disease severity score

In-hospital COVID-19 disease severity scores(12) were the sum of five components that range from 0 to 10 points, on admission: (1) age by decile (age greater than 60, 70, and 80 earned 1, 2, and 3 points, respectively), (2) mean arterial pressure (MAP) indicating hypotension (MAP below 80, 70, and 60 earned 1, 2, and 3 points), (3) oxygen saturation below 94% indicating impaired pulmonary function (1 point), (4) BUN greater than 30 indicating impaired renal function (1 point), and (5) INR greater than 1.2 and CRP greater than 10, indicating coagulopathy and inflammatory response. Scores ranged from 0 to 10 with higher score reflecting worse COVID-19 disease severity.

### Imaging assessment

Head computed tomography (CT) and brain magnetic resonance imaging (MRI) examinations were assessed at three different time points: those obtained before their COVID-19 hospitalization (most recent), during COVID-19 hospitalization, or after COVID-19 hospitalization (most recent) if available. Images were assessed by board-certified radiologists (K.H., J.L., each with at least 10 years of experience) and radiology residents (I.H., K.L., A.L.) under the supervision of the board-certified radiologists, blinded to the patient cohort designation. To establish assessment criteria and the scoring system, our board-certified radiologists and residents worked together to reach consensus by evaluating over a dozen images. Two residents scored each image and at least one board-certified radiologist reviewed and adjudicated. Major findings on both CT and MRI, documented for their absence or presence of hemorrhage, active stroke, prior stroke, mass effect, and microhemorrhage (on MRI only). In addition, white-matter (WM) change, microvascular disease (MVD), and volume loss on CT and MRI were graded as 0 for normal or not present, 1 for mild, 2 for moderate, and 3 for severe, taking account the *age* of patient. MRIs were additionally graded for extent of WM lesions or hyperintensities (HI) using the same grading scale. Findings were tabulated for pre-, during and post COVID-19. In cases where patients had both CT and MRI at a specific timepoint, MRI was used.

Finally, changes in imaging findings before and after COVID-19, if available, were assessed using the grading scale of 0: no change, ±1: mild change, ±2: moderate change, and ±3 severe change, with positive changes indicating worsening and negative indicating improvement between the two time points. Pre-COVID-19 images were used as a baseline if both pre- and during COVID-19 images were available.

### Primary outcomes

Primary outcomes were mortality, stroke, heart attack, MACE, reinfection, and hospital readmission post discharge January 23, 2023 (3 years post COVID-19). Secondary outcomes were qualitative and score-based clinical neuroimaging findings, which included the absence or presence of hemorrhage, active stroke, prior stroke, mass effect, and microhemorrhage, as well as scores of WM changes, MVD, and volume loss.

Associative models using logistic regression were employed to identify variables associated with mortality after discharge. Input for age was a continuous variable. Input for discharge disposition status was a single variable. Input for COVID-19 severity scores was a continuous variable. The rest of the variables used in the model were categorical variables. Odds ratios (OR) and 95% confidential interval (CI) were computed. In addition, Kaplan-Meier curves were constructed and analyzed using GraphPad Prism, with the outcome event being classified as dead (death date) or alive (most recent patient encounter). The survival curves of the neurological and control cohorts were compared with the logrank test (Mantel-Cox) method, resulting in a log rank hazard ratio (HR), 95% CI, and p-value.

### Statistical Analysis

Analysis of group differences of demographic and clinical variables employed χ^2^ tests for categorical variables and two-tailed t-tests for continuous variables via the statistical library in RStudio. Analysis of associative variables was performed in RStudio using a logistic regression model. Statistical comparison of imaging scores and changes of scores from baseline (pre or during COVID-19 hospitalization) to follow-up (post-COVID-19 hospitalization) employed unpaired t-test. P<0.05 was considered statistically significant unless otherwise specified. Statistics were not adjusted for multiple comparison due to the exploratory nature of the study.

### Standard Protocol Approvals, Registrations, and Patient Consents

This retrospective study using real world data was approved by our Institutional Review Board (#2021-13658) with a waiver of informed consent. All methods were performed in accordance with relevant guidelines and regulations pertaining to human subjects.

### Data Availability Statement

Reasonable request of data is available by contacting the corresponding author.

## RESULTS

Of the original neurological cohort of 636 patients, 414 were discharged alive and 371 returned to our health system. Of the original control cohort of 1743, 1199 were discharged alive and 1071 returned to our health system. The overall attrition rate was 10.60%.

### Discharge disposition

**Figure 1** shows the discharge dispositions of the neurological and control cohorts stratified by COVID-19 severity score. Patients with high severity scores were less likely to be discharged home and more likely to be discharged to a SNF or hospice in both groups. However, there were relatively more patients disposed to SNF and relatively fewer patients disposed to home in the neurological cohort compared to the control cohort.

**Figure 1.**
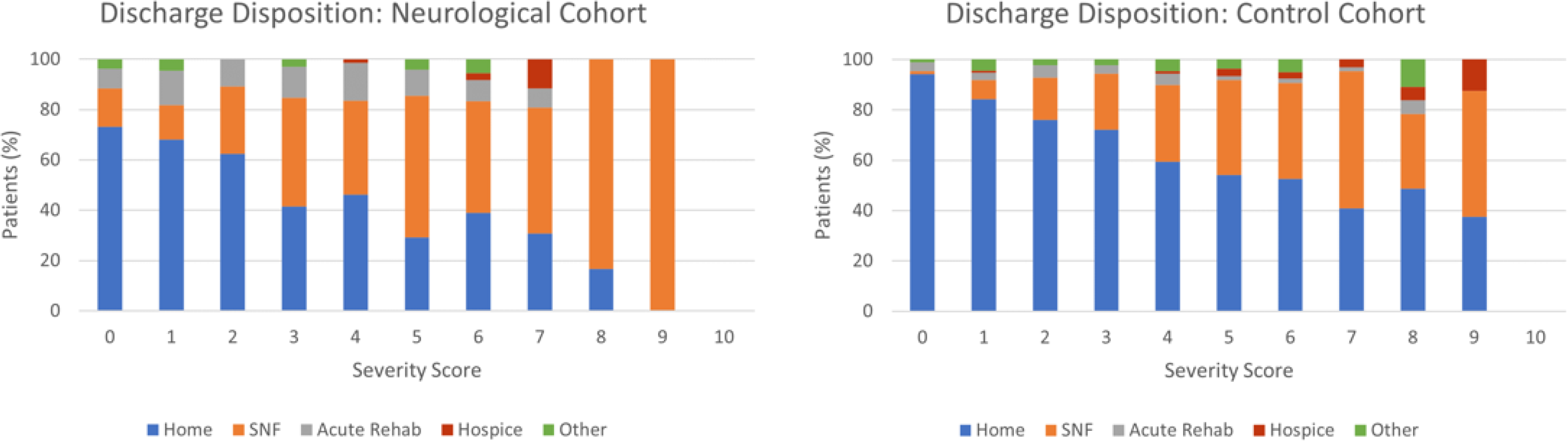
Percent of patients in the neurological and control cohorts discharged to home, acute rehabilitation, skilled nursing facility (SNF), hospice, and others for different COVID-19 severity score.

### Primary outcomes of neurological vs control cohort

**Table 1** shows the profiles of the survivors at discharge grouped by neurological and control cohorts. There were no significant group differences in age (69.71±15.80 vs 70.26±15.14, p>0.05), female gender composition (44.93% vs 48.21%, p>0.05), all major comorbidities (p>0.05) and race and ethnicity (p>0.05), except Black non-Hispanic (42.51% vs 36.03%, p=0.009), and Hispanic (33.57% vs 38.20%, p=0.047). More patients in the neurological cohort were discharged to acute rehabilitation (10.54% vs 3.68%, p<0.0001) and SNF (30.67% vs 20.78%, p=0.0002) and fewer survivors in the neurological cohort were discharged to home (55.27% vs 70.21%, p<0.0001) compared to the control cohort. With respect to laboratory data, there were few group differences between those at admission and at follow-up, as well as between groups (**Supplemental Table 1**). Some laboratory data at admission were worse than those at follow-up and laboratory data of the neurological cohort was worse than those of the control cohort.

**Table 1.** Demographics, comorbidities, and outcomes of survivor patients in the neurological and control cohorts. Mean±SD or n (%). * p<0.05, ** p<0.01, *** p<0.001 between the neurological and control cohorts.

|  | Neurologic Cohort<br>(N=414) | Control Cohort<br>(N=1199) |
| --- | --- | --- |
| **Patient Characteristics** |  |  |
| Age, years old (at admission) | 69.71±15.80 | 70.26±15.14 |
| Female (%) | 186 (44.93%) | 578 (48.21%) |
| **Combined Race and Ethnicity** |  |  |
| White, not Hispanic | 27 (6.52%) | 94 (7.84%) |
| Black, not Hispanic | 176 (42.51%) | 432 (36.03%)* |
| Hispanic | 139 (33.57%) | 458 (38.20%)* |
| Other | 62 (14.98%) | 169 (14.10%) |
| **Comorbidities (at admission)** |  |  |
| Hypertension | 186 (44.93%) | 523 (43.62%) |
| COPD/Asthma | 42 (10.14%) | 144 (12.01%) |
| Diabetes | 126 (30.43%) | 343 (28.61%) |
| CHF | 49 (11.84%) | 118 (9.84%) |
| CKD | 90 (21.74%) | 223 (18.60%) |
| **Discharge Disposition** |  |  |
| Home | 208 (50.24%) | 799 (66.64%)* |
| Acute Rehab | 43 (10.39%) | 40 (3.34%)* |
| SNF | 148 (35.75%) | 304 (25.35%)* |
| Hospice | 6 (1.45%) | 15 (1.25%) |
| Other | 9 (2.17%) | 41 (3.42%) |
| **Outcomes (Jan 10, 2023)** |  |  |
| Hospital Readmission | 272 (65.70%) | 728 (60.72%)* |
| Stroke | 26 (6.28%) | 28 (2.34%)* |
| Heart Attack | 22 (5.31%) | 50 (4.17%) |
| MACE | 85 (20.53%) | 198 (16.51%)* |
| **Mortality after discharge** |  |  |
| 0.5 years | 32 (7.73%) | 44 (3.67%)* |
| 1.0 years | 38 (9.18%) | 59 (4.92%)* |
| 3.0 years | 58 (14.01%) | 94 (7.84%)* |
| SARS-CoV-2 reinfection | 18 (4.35%) | 55 (4.59%) |

Incidence of readmission (65.70% vs 60.72%, p=0.036), stroke (6.28% vs 2.34%, p<0.0001), and MACE (20.53% vs 16.51%, p=0.032) were significantly higher in the neurological cohort than the control cohort. There were however no significant group differences in heart attack and reinfection after discharge (all p>0.05). Mortality rates were also higher in the neurological cohort compared to the control cohort at 0.5 years (7.73% vs 3.67%, p=0.0004), 1 year (9.18% vs 4.92%, p=0.0009), and 3 years (14.01% vs 7.84%, p=0.0001). Kaplan-Meier survival analysis (**Figure 2**) showed that the neurological cohort had a significantly lower survival probability than the control cohort at all timepoints (HR=2.346, 95% CI=(1.586, 3.470), p<0.0001).

**Figure 2.**
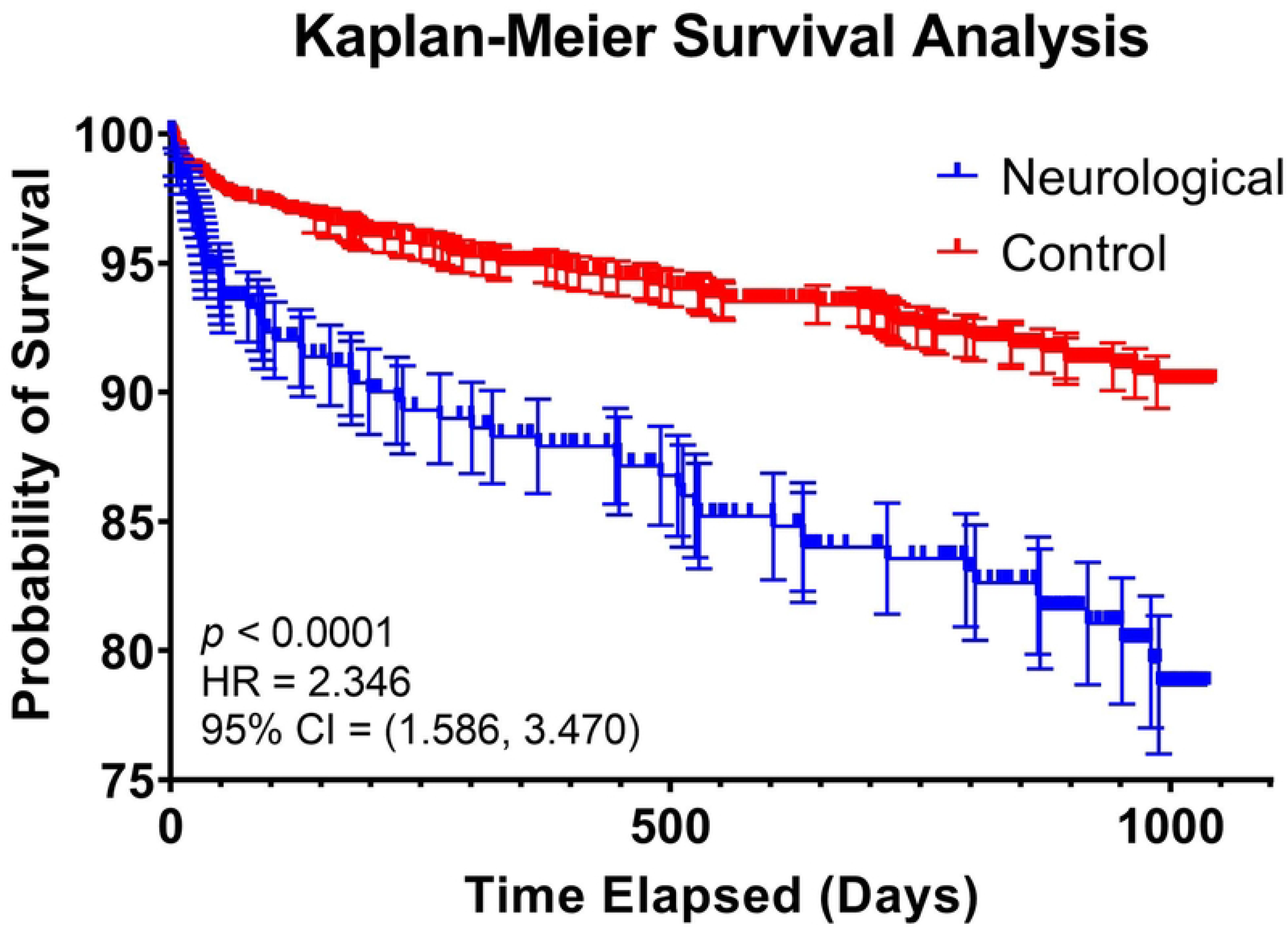
Kaplan-Meier Survival Curve analyzing the probability of survival after discharge from COVID-19 hospitalization in the neurological cohort versus the control cohort. HR = 2.346, 95% CI=(1.586, 3.470), p-value<0.0001.

### Outcomes versus COVID-19 severity scores

**Supplemental Figure 1A** shows the distribution of COVID-19 severity scores in each cohort, confirming proper propensity matching by severity score among survivors after COVID-19 hospitalization discharge. Primary outcomes were analyzed with respect to COVID-19 severity score for survivors and non-survivors post discharge (**Figure 1B-F**). Readmission for any medical reasons were similar among all severity scores for both cohorts. Incidence of stroke was high for all severity scores in the neurological cohort but was generally lower for matching scores in the control cohort. Incidence of heart attack also appeared to be trending upwards as severity score increased for both cohorts. The percent of patients who had MACE was distributed over a range of scores for both cohorts, with a higher COVID-19 severity scores showing a slightly higher percentage of patients with MACE at follow-up.

Non-survivors at follow-up had higher COVID-19 severity score compared to survivors.

### Cause of death post COVID-19 discharge

There were no group differences in cause of death between patients belonging to neurological and control cohorts (p>0.05). Beside the unknown cause of death (30.92%), the major causes of death after discharge were heart disease (13.79% neurological, 15.38% control), sepsis (8.62%, 17.58%), influenza and pneumonia (13.79%, 9.89%), COVID-19 (10.34%, 7.69%) and acute respiratory distress syndrome (ARDS) (10.34%, 6.59%) (**Table 2)**.

**Table 2.** Primary cause of death of patients who died after discharge. * p<0.05, ** p<0.01, *** p<0.001 between the neurological and control cohorts.

| **Cause of Death** | **Neurological**<br>**(N=58)** | **Control**<br>**(N=94)** |
| --- | --- | --- |
| Unknown | 16 (27.59%) | 31 (34.07%) |
| Heart Disease | 8 (13.79%) | 14 (15.38%) |
| Sepsis | 5 (8.62%) | 16 (17.58%) |
| Influenza and Pneumonia | 8 (13.79%) | 9 (9.89%) |
| COVID-19 | 6 (10.34%) | 7 (7.69%) |
| ARDS, Hypoxia | 6 (10.34%) | 6 (6.59%) |
| Other (GI Bleed, Pancreatitis, ICH, Acute Liver Failure) | 3 (5.17%) | 6 (6.59%) |
| Cancer | 2 (3.45%) | 2 (2.20%) |
| Nephritis, Nephrotic Syndrome, and Nephrosis | 2 (3.45%) | 1 (1.10%) |
| Stroke (Cerebrovascular Diseases) | 1 (1.72%) | 1 (1.10%) |
| Accidents (Unintentional Injuries) | 1 (1.72%) | 1 (1.10%) |

### Survivors and non-survivors post discharge

**Table 3A** compared the profiles of survivors and non-survivors in the neurological and control cohorts post discharge. There are few differences of survivors and non-survivors between cohorts. Both within the neurological and control cohorts, non-survivors were significantly older and had higher COVID-19 severity score compared to survivors. Although no differences in comorbidities were observed in the neurological cohort, control non-survivors had higher incidence of hypertension, diabetes, CHF, and CKD. Both neurological and control non-survivors were less likely to be discharged home and more likely to be discharged to SNF.

**Table 3.** (A) Demographics and comorbidities of patients who died versus survived after discharge in the neurological and control cohorts. Patients lost to follow up were excluded. Mean±SD or n (%). * p<0.05, ** p<0.01, *** p<0.001 between survivors or non-survivors in the neurological and control cohorts. ^ p<0.05, ^^ p<0.01, ^^^ p<0.001 between survivors and non-survivors in the same cohort. (B) Odds ratios of mortality post discharge.

| (A) | Neurological Cohort (N=371) |  | Control Cohort (N=1043) |  |
| --- | --- | --- | --- | --- |
|  | Survivors<br>N=313 (84.37%) | Non-survivors<br>N=58 (15.63%) | Survivors<br>N=949 (90.99%) | Non-survivors<br>N=94 (9.01%) |
| **Patient Characteristics** |  |  |  |  |
| Age, year old (at admission) | 67.85±15.85 | 76.40±11.71*** | 69.20±14.95 | 76.56±11.23*** |
| Female | 147 (46.96%) | 22 (37.93%) | 480 (50.58%) | 43 (45.74%) |
| Combined Race and Ethnicity |  |  |  |  |
| White, non-Hispanic | 23 (7.35%) | 3 (5.17%) | 68 (7.17%) | 9 (9.57%) |
| Black, non-Hispanic | 129 (41.21%) | 28 (48.28%) | 350 (36.88%) | 42 (44.68%) |
| Hispanic | 106 (33.87%) | 19 (32.76%) | 383 (40.36%) | 39 (41.49%) |
| Other | 47 (12.02%) | 5 (8.62%) | 139 (14.65%) | 3 (3.19%)* |
| COVID-19 Severity Score | 3.32±2.02 | 4.40±1.79 | 3.36±2.06^ | 4.74±2.05***, ^^ |
| **Comorbidities (at admission)** |  |  |  |  |
| Hypertension | 151 (48.24%) | 31 (53.45%) | 456 (48.05%) | 54 (57.45%)* |
| COPD/Asthma | 35 (11.18%) | 6 (10.34%) | 128 (13.49%) | 17 (18.09%) |
| Diabetes | 100 (31.95%) | 21 (36.21%) | 298 (31.40%) | 39 (41.49%)* |
| Congestive heart failure | 38 (12.14%) | 11 (18.97%) | 83 (8.75%) | 31 (32.98%)*, ^^ |
| Chronic kidney disease | 75 (23.96%) | 14 (24.14%) | 185 (19.49%) | 36 (38.30%)* |
| **Discharge Disposition** |  |  |  |  |
| Home | 173 (55.27%) | 17 (29.31%)* | 686 (72.29%) | 38 (40.43%)* |
| Acute Rehab | 33 (10.54%) | 7 (12.07%) | 36 (3.79%) | 4 (4.26%) |
| SNF | 96 (30.67%) | 32 (55.17%)* | 203 (21.39%) | 39 (41.49%)* |
| Hospice | 2 (0.64%) | 2 (3.45%) | 1 (0.11%) | 4 (4.26%)* |
| Other | 9 (2.88%) | 0 (0.00%) | 23 (2.42%) | 9 (9.57%)* |

| (B) | OR | 95% CI | p-val |
| --- | --- | --- | --- |
| Belonging to Neurological Cohort | 1.802 | (1.237, 2.608) | **0.002** |
| Discharge disposition | 1.508 | (1.276, 1.775) | **<0.0001** |
| Congestive heart failure | 2.281 | (1.429, 3.593) | **0.0004** |
| COVID-19 Severity Score | 1.177 | (1.062, 1.304) | **0.002** |
| Age | 1.027 | (1.010, 1.044) | **0.002** |
| Male | 1.387 | (0.963, 2.004) | 0.080 |
| Chronic kidney disease | 1.321 | (0.849, 2.039) | 0.213 |
| Diabetes | 1.131 | (0.744, 1.710) | 0.561 |
| COPD/Asthma | 1.130 | (0.651, 1.892) | 0.652 |
| Hypertension | 0.985 | (0.649, 1.492) | 0.942 |

### Risk factors for mortality after discharge

A logistic regression model found 5 significant variables associated with mortality post discharge (**Table 3B**). These variables were belonging to the neurological cohort (OR=1.802 (1.237, 2.608), p=0.002), discharge disposition (OR=1.508, 95% CI=(1.276, 1.775), p<0.0001), congestive heart failure (OR=2.281 (1.429, 3.593), p=0.0004), COVID-19 severity score (OR=1.177 (1.062, 1.304), p=0.002), and age (OR=1.027 (1.010, 1.044), p=0.002). Male gender (OR=1.387 (0.963, 2.004), p=0.08) was trending significance.

### Imaging findings

**Table 4A** summarizes the pre-, intra-, and post- COVID-19 neuroradiological findings. The numbers of pre-, intra-, and post- COVID-19 patients with imaging studies varied. Of those with imaging, about 20% were MRI and 80% were CT. For qualitative assessment, 30-40% of patients had prior strokes for all three time points, whereas the presence of hemorrhage, active stroke and/or mass effect were relatively low (0-10% with most around 5%). There were, however, no group differences in these qualitative findings for all three time points (p>0.05).

**Table 4.** Imaging findings pre, during, and post-COVID-19 infection. The neurological cohort had pre- and post-COVID-19 imaging at 456±729 days and 335±274 days before and after hospitalization, respectively. The control cohort had pre- and post-COVID-19 imaging at 828±974 and 432±300 days before and after hospitalization, respectively. Patients who died during COVID-19 hospitalization are excluded. Studies were scored as 0 (normal or no abnormality), 1 (mild abnormality), 2 (moderate abnormality), and 3 (severe abnormality).

|  | Pre COVID-19 |  |  | During COVID-19 |  |  | Post COVID-19 |  |  |
| --- | --- | --- | --- | --- | --- | --- | --- | --- | --- |
|  | Neurological | Control | p-val | Neurological | Control | p-val | Neurological | Control | p-val |
| Total Patients | 94 | 143 |  | 65 | 32 |  | 144 | 251 |  |
| MRI | 21 (22.34%) | 32 (22.38%) | 1.000 | 6 (9.23%) | 6 (18.75%) | 0.312 | 32 (21.92%) | 67 (26.69%) | 0.386 |
| CT | 73 (77.66%) | 111 (77.62%) | 1.000 | 59 (90.77%) | 26 (81.25%) | 0.312 | 112 (76.71%) | 184 (73.31%) | 0.386 |
| **Qualitative Findings** |  |  |  |  |  |  |  |  |  |
| Hemorrhage | 4 (4.26%) | 3 (2.10%) | 0.570 | 5 (7.69%) | 0 (0%) | 0.262 | 7 (4.86%) | 7 (2.79%) | 0.430 |
| Active Stroke | 6 (6.38%) | 7 (4.90%) | 0.841 | 7 (10.77%) | 2 (6.25%) | 0.727 | 11 (7.64%) | 12 (4.78%) | 0.345 |
| Prior Stroke | 35 (37.23%) | 43 (30.07%) | 0.314 | 23 (35.38%) | 11 (34.38%) | 1.000 | 58 (40.28%) | 76 (30.28%) | 0.056 |
| Mass Effect | 2 (2.13%) | 7 (4.90%) | 0.457 | 6 (9.23%) | 2 (6.25%) | 0.913 | 10 (6.94%) | 14 (5.58%) | 0.743 |
| Microhemorrhage | 5 (5.32%) | 8 (5.59%) | 1.000 | 5 (4.62%) | 0 (0%) | 0.262 | 9 (6.25%) | 17 (6.77%) | 1.000 |
| **Scoring Findings** |  |  |  |  |  |  |  |  |  |
| White Matter Change | 0.99±0.81 | 0.93±0.75 | 0.572 | 0.97±0.78 | 1.13±0.89 | 0.411 | 1.06±0.86 | 1.05±0.84 | 0.942 |
| 0 | 27 (28.72%) | 41 (28.67%) | 1.000 | 18 (27.69%) | 7 (21.88%) | 0.712 | 39 (27.08%) | 66 (26.69%) | 0.958 |
| 1 | 45 (47.87%) | 76 (53.15%) | 0.508 | 34 (52.31%) | 18 (56.25%) | 0.881 | 67 (46.53%) | 122 (48.61%) | 0.769 |
| 2 | 18 (19.15%) | 21 (14.69%) | 0.467 | 10 (15.38%) | 3 (9.38%) | 0.617 | 28 (19.44%) | 47 (18.73%) | 0.966 |
| 3 | 4 (4.26%) | 5 (3.50%) | 1.000 | 3 (4.62%) | 4 (12.50%) | 0.320 | 10 (6.94%) | 16 (6.37%) | 0.993 |
| Microvascular Disease | 0.95±0.78 | 0.89±0.75 | 0.566 | 0.94±0.80 | 1.16±0.87 | 0.246 | 1.05±0.84 | 1.04±0.83 | 0.922 |
| 0 | 28 (29.79%) | 45 (31.47%) | 0.896 | 20 (30.77%) | 6 (18.75%) | 0.311 | 39 (27.08%) | 67 (26.69%) | 1.000 |
| 1 | 46 (48.94%) | 73 (51.05%) | 0.853 | 32 (49.23%) | 19 (59.38%) | 0.469 | 68 (47.22%) | 123 (49.00%) | 0.813 |
| 2 | 17 (18.09%) | 21 (14.69%) | 0.605 | 10 (15.38%) | 3 (9.38%) | 0.617 | 28 (19.44%) | 46 (18.33%) | 0.889 |
| 3 | 3 (3.19%) | 4 (2.80%) | 1.000 | 3 (4.62%) | 4 (12.50%) | 0.320 | 9 (6.25%) | 15 (5.98%) | 1.000 |
| Volume Loss | 0.55±0.68 | 0.48±0.59 | 0.412 | 0.63±0.67 | 0.66±0.77 | 0.876 | 0.72±0.71 | 0.57±0.69 | **0.045** |
| 0 | 51 (54.26%) | 81 (56.64%) | 0.819 | 31 (47.69%) | 16 (50.00%) | 1.000 | 61 (42.36%) | 135 (53.78%) | **0.037** |
| 1 | 35 (37.23%) | 55 (38.46%) | 0.957 | 27 (41.54%) | 12 (37.50%) | 0.872 | 63 (43.75%) | 89 (35.46%) | 0.128 |
| 2 | 7 (7.45%) | 7 (4.90%) | 0.594 | 7 (10.77%) | 3 (9.38%) | 1.000 | 19 (13.19%) | 26 (10.36%) | 0.491 |
| 3 | 1 (1.06%) | 0 (0%) | 0.832 | 0 (0%) | 1 (3.13%) | 0.716 | 1 (0.69%) | 1 (0.40%) | 1.000 |
| White Matter Hyperintensity/Lesion | 0.62±0.50 | 0.38±0.28 | 0.338 | 1±0.38 | 0.33±0.24 | 0.645 | 0.41±0.39 | 0.49±0.43 | 0.333 |
| 0 | 13 (13.83%) | 20 (13.99%) | 1.000 | 2 (3.08%) | 4 (12.50%) | 0.173 | 23 (15.97%) | 41 (16.33%) | 1.000 |
| 1 | 4 (4.26%) | 12 (8.39%) | 0.329 | 2 (3.08%) | 2 (6.25%) | 0.845 | 6 (4.17%) | 21 (8.37%) | 0.166 |
| 2 | 3 (3.19%) | 0 (0%) | 0.120 | 2 (3.08%) | 0 (0%) | 0.808 | 2 (1.39%) | 3 (1.20%) | 1.000 |
| 3 | 1 (1.06%) | 0 (0%) | 0.832 | 0 (0%) | 0 (0%) | NA | 1 (0.69%) | 2 (0.80%) | 1.000 |

For score-based assessment, the average scores for age-appropriate WM change and MVD were about 1 (mild abnormality), and the average scores for age-appropriate volume loss and WM lesions were about 0.5 (no to mild abnormality). Distribution of scores were similar between groups. There were no differences in scores between groups, except for volume loss post COVID-19 (average score: 0.72±0.71 neurological vs 0.57±0.69 control, p=0.045; and score of 0: 42.36% neurological vs. 53.78%, p=0.037).

**Table 4B** shows the changes in imaging findings between baseline (pre- or intra-COVID-19) and follow-up (post-COVID-19). There were significant increases in the incidence of active stroke (neurological: 6.25%, p=0.021; control: 3.82%, p=0.039), prior stroke (12.5%, p=0.0003; 9.55%, p=0.002), and microhemorrhage (7.14%, p=0.012; 6.37%, p=0.004) between the two time points, indicative of age-related effects. There were, however, no group differences (p>0.05 for all).

**Table 4B.** Changes in imaging findings between baseline (pre or during COVID-19 hospitalization) and follow-up (post-COVID-19 hospitalization). Only patients with both a baseline and follow-up scan were included. Studies were scored as 0 (normal or no abnormality), 1 (mild abnormality), 2 (moderate abnormality), and 3 (severe abnormality).

|  | Neurological (N=112) |  | Control (N=157) |  |  |
| --- | --- | --- | --- | --- | --- |
|  |  | p-val |  | p-val | p-val (neurological vs control) |
| **Changes in incidence for** |  |  |  |  |  |
| Hemorrhage | 3 (2.68%) | 0.245 | 3 (1.91%) | 0.246 | 0.999 |
| Active Stroke | 7 (6.25%) | **0.021** | 6 (3.82%) | **0.039** | 0.531 |
| Prior Stroke | 14 | **0.0003** | 15 (9.55%) | **0.0002** | 0.570 |
| Mass Effect | 5 (4.46%) | 0.070 | 4 (2.55%) | 0.131 | 0.605 |
| Microhemorrhage | 8 (7.14%) | **0.012** | 10 (6.37%) | **0.004** | 0.998 |
| **Changes in scores for** |  |  |  |  |  |
| White Matter Change | 0.12±0.44 | **0.006** | 0.18±0.46 | **<0.0001** | 0.219 |
| Microvascular Disease | 0.15±0.41 | **0.0001** | 0.18±0.46 | **<0.0001** | 0.538 |
| Volume Loss | 0.15±0.41 | **0.0001** | 0.17±0.42 | **<0.0001** | 0.787 |
| White Matter Hyperintensity/Lesion | -0.04±0.63 | 0.459 | 0.03±0.42 | 0.452 | 0.310 |

WM changes (neurological: 0.12±0.44, p=0.006; control: 0.18±0.46, p<0.0001), MVD (0.15±0.41, p=0.0001; 0.18±0.46, p<0.0001) and volume loss (0.15±0.41, p=0.0001; 0.17±0.42, p<0.00101) were higher (worsening) post-COVID compared to baseline for both neurological and control cohorts. There were, however, also no group differences (p>0.05 for all).

## DISCUSSION

This study investigated the 3-year outcomes of hospitalized COVID-19 patients with and without major neurological issues. The major findings are: i) COVID-19 patients with significant neurological issues that warranted neuroimaging were more likely to be discharged to acute rehabilitation and skilled nursing facilities compared to matched controls, ii) the neurological cohort had higher mortality rates after discharge (HR=2.346, p<0.0001) compared to controls, iii) the incidence of readmission, stroke, and MACE, but not heart attack or reinfection, were higher in the neurological cohort at 3 years follow-up, iv) the primary causes of death after discharge for both cohorts were heart failure, sepsis, influenza and pneumonia, COVID-19 and ARDS, v) patients who died post-discharge were significantly older, more likely to be male, had higher COVID-19 severity score, and those were sent to skilled nursing facilities at discharge compared to survivors, vi) there were no group differences in gross radiological findings with respect to hemorrhage and stroke, except the neurological cohort showed significantly more age-appropriate volume loss than the control cohort.

### Disposition

Approximately half of the neurological patients and one-thirds of the control patients were discharged to SNF, acute rehabilitation, or hospice. These findings indicated that many COVID-19 patients were not functionally independent after discharge(22–24), especially those in the neurological cohort. Few studies today have reported home, SNF, and acute rehabilitation discharge rates after COVID-19 hospitalization(22–26). These findings suggest that patients in the neurological cohort likely needed more follow-up medical care at discharge.

### Primary outcomes

The majority (60-65%) of all neurological and control patients were readmitted to our health system for medical reasons over 3 years. This is not surprising for our cohort due to advanced age and high prevalence of comorbidities, although it is higher than reported in some studies(27–30).

Readmission could be due to age-related illness or medical conditions exacerbated by COVID-19. The neurological cohort had a higher readmission rate than the control cohort, suggesting that a higher burden of disease during COVID-19 hospitalization may be associated with a higher probability of re- hospitalization.

The incidence of stroke was 2-6% and of heart attack was 4-5% in both groups. The incidence of MACE after discharge (16-20%) was higher than other reported previously in COVID-19 who did not have neurological issues(18). A few studies have suggested COVID-19 exerts long-term cardiovascular effects(16, 31–33), consistent with a disease that affects the cardiovascular system and thus can result in MACE after severe infection warranting hospitalization. Approximately 4-5% of both cohorts experienced COVID-19 re-infection. This rate of re-infection is slightly higher than what is reported elsewhere(34, 35). The higher rate of re-infection may be attributed to the urban setting of congested environs, high rates of comorbidities and healthcare disparities associated with lower socioeconomic status(36).

Of those who died post-discharge, more than half died within the first 0.5 years in both groups, suggesting that most of these deaths were likely COVID-19 related. The cumulative mortality rates of the neurological and control cohorts at 3 years post-discharge were 14% and 8%, respectively. The marked mortality rate differences between groups are highlighted by the Kaplan-Meier analysis. Those who died at follow-up in both groups were 8 years older and more likely to be male as compared to survivors.

Elderly patients may be more prone to exhibiting early neurologic symptoms because of limited cognitive reserve. This is widely seen in other medical conditions such as urosepsis. Patients presenting with early neurologic compromise could be a harbinger of susceptibility for higher mortality across other disease states. Male gender has been previously reported to have worse acute in-hospital outcomes, including higher rate of multi-organ injury, critical care illness, and in-hospital mortality(37–40). Here we reported male gender also had worse long-term outcomes post COVID-19 discharge.

Logistic regression model identified discharge disposition to be the top risk factor for post-discharge mortality, followed by CHF, COVID-19 severity score, belonging to the neurological cohort, and age. CHF was the only comorbidity that was significantly associated with post-discharge mortality. Patients who had more severe COVID-19 disease were also more likely to die post discharge. Belonging to the neurological cohort was also independent risk factor for post-discharge mortality. It is not surprising that advanced age is associated with higher post-discharge mortality, but advanced age ranked lower than other variables mentioned above. Note that OR for male gender was trending significance and we predicted that large sample sizes could result in significant findings. Taken together, these findings underscore the independent risk factors that contributed to post discharge mortality and notably belonging to the neurological cohort is a significant independent risk factor.

### Cause of Death

The causes of death were similar between neurological and control cohorts, consistent with findings using logistic regression in which belonging to neurological cohort was an important but not the most important associative variable of post COVID-19 discharge mortality. The primary known causes of death in both the neurological and control cohorts were heart disease, sepsis, influenza and pneumonia, COVID-19, and ARDS. Sepsis, pneumonia, and ARDS might be related to or be triggered by COVID-19, although they could also be a result of other medical events. It is possible that COVID-19 as a cause of death was underestimated as a result of imprecise categorization in the death certificates. Note that about one-third of the causes of death were specified as unknown on the death certificates. It is possible that some patients died of senescence and no primary cause of death was noted.

### Imaging Findings

Age of patient was taken into consideration when assessing neuroradiological findings. Imaging findings of control and neurological patients displayed differential profiles of abnormalities that were consistent with age and comorbidities in this population. The differences in radiological findings between baseline and follow-up showed age-related effects. However, there were generally no group differences in qualitative nor score-based findings, except that the neurological cohort showing higher volume loss post-COVID-19 compared to controls.

Several case reports and few cohort studies have identified reduction in grey-matter thickness, ischemic stroke, decrease in global brain size, cerebral microstructural changes, and persistent WM changes associated with COVID-19(41–47). There is likely some reporting bias in case or case-series studies as positive clinical imaging findings associated with COVID-19 are more likely to be reported. Most of these studies do not compare findings to baseline, which makes it difficult to discern whether imaging abnormalities were pre-existing or a consequence of COVID-19 disease. None of these studies employ a scoring system to accentuate the degree of abnormality. Our study is novel because of its large and diverse patient population, long follow-up times, the use of a scoring system, and comparison between baseline and follow-up studies to 3 years post-discharge. It is possible that COVID-19 related changes in the brain anatomy and structure could take time to manifest, and we predict that some COVID- 19 patients will likely experience accelerated aging and high incidence of age-related disorders. Brain imaging is important because it could provide neural correlates of post-COVID-19 neurological sequela, which include, but is not limited to, neurological symptoms, neurocognitive symptoms, fatigue, memory loss, anxiety, depression, and post-traumatic stress disorder(8, 48–50).

Taken together these current observations contribute new insights concerning our understanding of the longitudinal effects of neurological involvement in long COVID. In terms of nervous system involvement, persistence and evolution, there are likely bidirectional interactions between the nervous and the immune systems that orchestrate a composite pro-inflammatory, hypercoagulable, hypoxemic, and immune dysregulated state(7). In addition, these dynamic processes could contribute to accelerated brain aging, stress pathway-mediated neural injury responses, features of traumatic encephalopathy, demyelination, neurodegeneration, and accompanying preferential cortical atrophy as we have identified in this study. Moreover, multiple communication routes between the central and peripheral nervous systems and the body in long COVID create systemic organ system, tissue and cellular interfaces that impair organismal homeostasis to give rise to persistent deregulated regenerative and plasticity responses. These pathological processes promote chronic multi-organ dysfunction and can lead to a spectrum of stressor states that predispose to organ fibrosis, tissue degeneration and even dysplastic and neoplastic conditions with associated metabolic derangement, immune dysregulation, inflammatory processes, and additional features of SARS-CoV-2/COVID-19-mediated dyshomeostasis syndrome, including proteotoxicity and protean epigenetic derangements. Such biological contingencies suggest that the multifactorial nature of the neurological manifestations of COVID-19 may put patients at higher risk of long-term functional disabilities and death as we have currently observed. Importantly, our increasing understanding of the nature of the deregulation of dynamic nervous system-systemic crosstalk displayed in response to SARS-CoV-2 may allow us to devise innovative and interdisciplinary mitigation strategies to alleviate the long-term sequelae preferentially caused by early neurological involvement in COVID-19.

### Limitations

This is one of the largest cohort studies of imaging findings and one of the longest follow-up studies of COVID-19 survivors. This study however has several limitations. Our patient cohort was limited to patients infected with COVID-19 during the first wave of the pandemic, when hospitals were overburdened, COVID-19 vaccines were not yet available, and COVID-19 treatments were limited. The patient profiles (i.e., age composition) might differ from those of subsequent waves. Additionally, the selection of patients was subjective and may have introduced selection bias. In building the control cohort, propensity scoring only factored in age and COVID-19 severity to remain consistent with the index study.

Although the attrition is low (10%), patients who did not return to our health system could not be studied. While it is possible that returning patients were more likely to have more severe COVID-19, our patient data consisted of those who returned for any medical reasons, including regular checkups. On the other hand, those who did not return might have expired. Our current study was not powered to address differences due to race and ethnicity because our cohort consisted of large proportions of Blacks and Hispanics but low proportions of other race and ethnic groups. Imaging sample sizes were small because not all patients had imaging performed at all three time points. The mixture of MRI and CT, which may have different sensitivities for various accompanying pathologies. More sophisticated imaging methods are warranted. Future studies should compare results with those of the general population without SARS-CoV-2 infection. Other factors such as reinfection, COVID-19 vaccination status, and influenza vaccination status could affect long-term outcomes. As with any retrospective study, there could be other unintended patient selection biases and unaccounted for confounders.

## Conclusions

Patients with significant neurological findings during COVID-19 hospitalization were more likely to have worse outcomes at 3-year follow-up compared to propensity matched controls. Improved understanding of the long-term outcomes of COVID-19 patients with neurological involvement could help to develop effective screening methods and innovative interventions to address the potentially high burden of care among these COVID-19 survivors.

## Data Availability

Data available by contacting the corresponding author.

## Acknowledgments

None

## ABBREVIATIONS

ALT: alanine aminotransferase
ARDS: acute respiratory distress syndrome
AST: aspartate transaminase
BNP: brain natriuretic peptide
BUN: blood urea nitrogen
CDM: Common Data Model
CHF: congestive heart failure
CI: confidence interval
CKD: chronic kidney disease
CNS: central nervous system
COPD: chronic obstructive pulmonary disease
CRP: C-reaction protein
CT: computed tomography
Cr: creatinine
DDIM: D-dimer
EMR: electronic medical record
HI: hyperintensity
HR: hazard ratio
INR: international normalized ratio
LDH: lactate dehydrogenase
Lymph: lymphocyte count
MACE: major adverse cardiovascular events
MAP: mean arterial pressure
MRI: magnetic resonance imaging
MVD: microvascular disease
OHDSI: Observational Health Data Sciences and Information
OMOP: Observational Medical Outcomes Partnership
OR: odds ratio
SBP: systolic blood pressure
SNF: skilled nursing facility
TNT: troponin-T
WBC: white blood cell count
WM: white matter

---

**Supplemental Table 1.** Laboratory values of neurological and control patients at COVID-19 hospitalization admission and most recent follow-up. Mean±SD or n (%). * p<0.05, ** p<0.01, *** p<0.001 between the neurological and control cohorts. ^ p<0.05, ^^ p<0.01, ^^^ p<0.001 between admission and follow-up lab values within the same cohort.

| Lab Values | Neurologic Cohort<br>(N=414) |  | Control Cohort<br>(N=1199) |  |
| --- | --- | --- | --- | --- |
|  | Admission | Follow-Up | Admission | Follow-Up |
| Temperature [°F] | 98.86±1.22<sup>^^</sup> | 98.22±0.74 | 99.05±1.33<sup>^^^*</sup> | 98.11±2.05 |
| SBP [mmHg] | 133.64±27.46<sup>^^</sup> | 124.19±24.44 | 135.09±25.25<sup>^^</sup> | 127.24±22.49* |
| Mean arterial pressure [mmHg] | 122.38±83.16 | 116.77±61.21 | 103.33±65.24<sup>^^^***</sup> | 111.56±63.52 |
| DDIM [ug/mL FEU] | 4.27±5.57<sup>^^</sup> | 3.17±4.42 | 3.46±4.84<sup>**</sup> | 3.05±14.05 |
| INR [seconds] | 15.75±7.00 | 15.48±4.48 | 15.32±6.82 | 15.38±4.99 |
| BUN [mg/dL] | 35.35±35.08<sup>^^</sup> | 26.26±24.06 | 29.94±28.38<sup>^^^**</sup> | 22.64±20.52<sup>**</sup> |
| Cr [mg/dL] | 2.15±2.74<sup>^</sup> | 1.79±2.27 | 1.90±2.43<sup>^^</sup> | 1.57±1.90* |
| Sodium | 138.58±8.07 | 139.18±4.85 | 137.75±7.48<sup>^^</sup> | 139.10±4.30 |
| Glucose [mg/dL] | 190.21±179.35<sup>^^</sup> | 136.47±74.92 | 172.68±135.56<sup>^^^*</sup> | 130.12±62.67 |
| AST [U/L] | 48.76±53.08 | 61.20±257.39 | 56.87±105.55* | 52.89±361.85 |
| ALT [U/L] | 33.85±44.55 | 51.74±267.41 | 38.82±71.60* | 36.96±102.81 |
| WBC [k/uL] | 8.13±4.67 | 7.95±3.99 | 8.47±7.31<sup>^^</sup> | 7.91±3.66 |
| Lymph [k/uL] | 1.16±0.76<sup>^^</sup> | 1.67±0.93 | 1.38±4.94<sup>^^</sup> | 1.74±1.09 |
| FERR [ng/mL] | 1251.76±1550.73<sup>^</sup> | 964.16±2385.43 | 1261.31±2092.20<sup>^^</sup> | 948.88±1532.12 |
| CRP [mg/L] | 9.61±9.33<sup>^^</sup> | 5.91±6.94 | 11.10±9.66<sup>^^^**</sup> | 5.54±6.27 |
| Procalcitonin [ug/L] | 1.99±6.17 | 2.23±7.15 | 1.25±4.82* | 1.43±6.10* |
| LDH [U/L] | 411.72±300.88<sup>^^</sup> | 345.73±156.67 | 404.41±231.39<sup>^^</sup> | 341.15±267.74 |
| BNP [pg/mL] | 2519.11±4491.13<sup>^^</sup> | 1642.03±3577.47 | 1825.85±3716.69<sup>^^^*,*</sup> | 1242.17±2948.39* |
| TNT [ug/L] | 0.06±0.19<sup>^</sup> | 0.26±2.13 | 0.04±0.09<sup>^^^*</sup> | 0.18±1.53 |
| Pulse Oximetry [% Oxygenation] | 95.20±5.51<sup>^^</sup> | 97.10±6.31 | 93.80±7.15<sup>^^^,***</sup> | 97.12±5.56 |

**Supplemental Figure 1.**
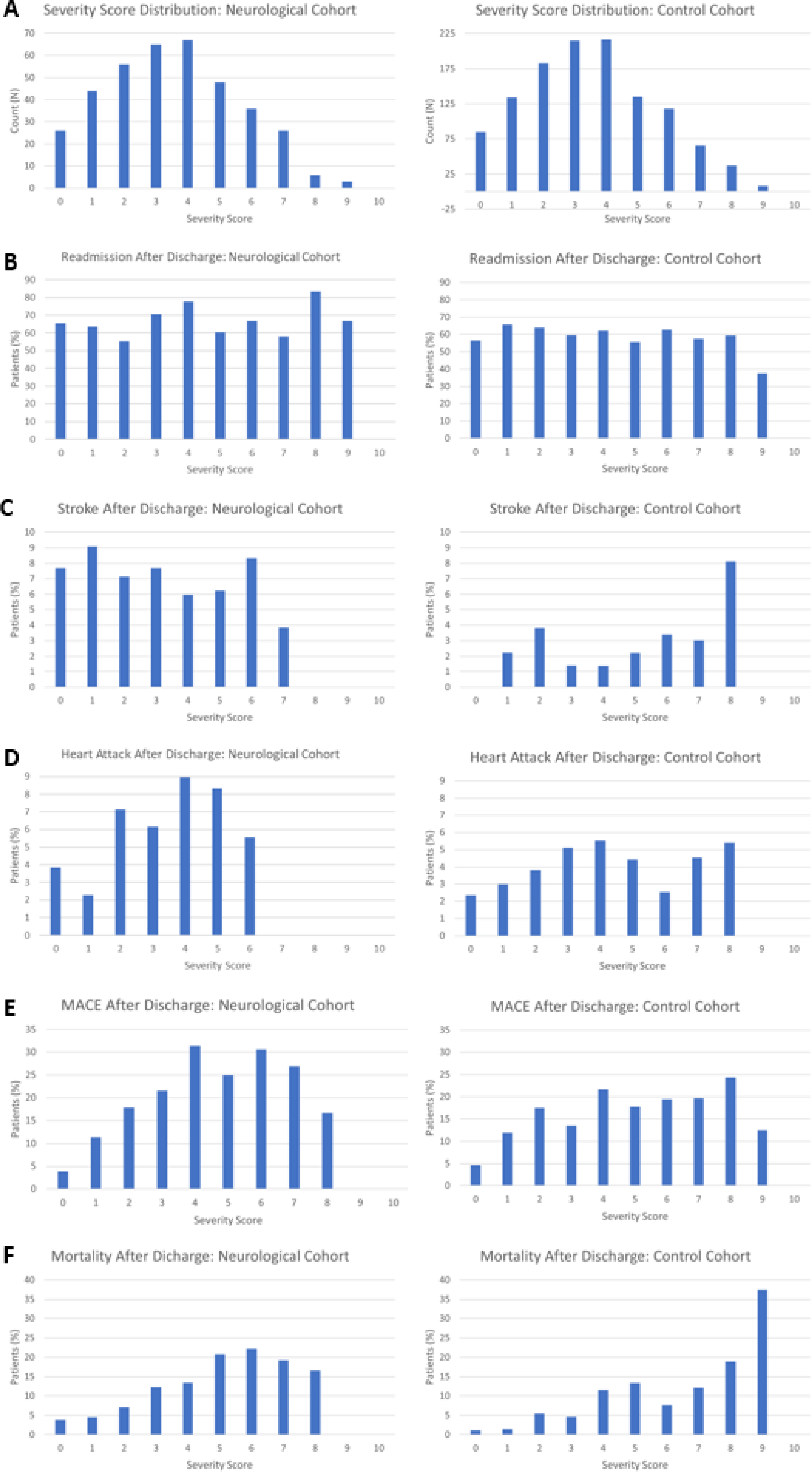
(A) Distribution of COVID-19 severity score in the neurological and control cohorts (survivors after COVID-19 hospitalization discharge). Percent of patients in the neurological and control cohorts who (B) were readmitted, (C) had stroke, (D) had heart attack, (E) had MACE, and (F) died after discharge from COVID-19 hospitalization.

